# Cancer predisposition syndromes as risk factor for early second primary neoplasms after childhood cancer – a national cohort study

**DOI:** 10.1101/2020.09.05.20180612

**Authors:** Nicolas Waespe, Fabiën N. Belle, Shelagh Redmond, Christina Schindera, Ben D. Spycher, Jochen Rössler, Marc Ansari, Claudia E. Kuehni, for the Swiss Pediatric Oncology Group (SPOG)

## Abstract

**Background:** Childhood cancer patients are at increased risk of second primary neoplasms (SPNs). We assessed incidence and risk factors for early SPNs with a focus on cancer predisposition syndromes (CPSs).

**Patients and methods:** This cohort study used data from the Swiss Childhood Cancer Registry. We included patients with first primary neoplasms (FPN) until age 21 years from 1986 to 2015 and identified SPNs occurring before age 21. We calculated standardized incidence ratios (SIR) and absolute excess risks (AER) using Swiss population cancer incidence data and cumulative incidence of SPNs. We calculated hazard ratios (HR) of risk factors for SPNs using Fine and Gray competing risk regression.

**Results:** Among 8,074 childhood cancer patients, 304 (4%) were diagnosed with a CPS and 94 (1%) developed early SPNs. The incidence of SPNs was more than 10-fold increased in childhood cancer patients compared to neoplasms in the general population (SIR 10.6, 95%-confidence interval [CI] 8.7-13.1) and the AER was 179/100,000 person-years (CI 139-219). Cumulative incidence of SPNs 20 years after FPN diagnosis was 23% in patients with CPSs and 3% in those without. Risk factors for SPNs were CPSs (HR 7.8, CI 4.8-12.7), chemotherapy (HR 2.2, CI 1.1-4.6), radiotherapy (HR 1.9, CI 1.2-2.9), hematopoietic stem cell transplantation (HR 1.8, CI 1-3.3), and older age (15-20 years) at FPN diagnosis (HR 1.9, CI 1.1-3.2).

**Conclusion:** CPSs are associated with a high risk of SPNs before age 21 years. Identification of CPSs is important for appropriate cancer surveillance and targeted screening.

## 1. Introduction

While most children can be cured of their first primary neoplasm (FPN) with modern treatments, they remain at risk of second primary neoplasms (SPNs) throughout their life. SPNs are an important cause of morbidity and the most common cause of death in long-term childhood cancer survivors.[1-4] Early SPNs, defined here as occurring under 21 years of age, have been mostly studied together with SPNs occurring later in life. From many studies, early SPNs were excluded because the analysis had been restricted to adult survivors.[2,5-9] The incidence ratio of neoplasms, when comparing cancer patients to the general population, is highest during childhood and adolescence.[10-12] Overall survival of patients with SPNs is low and poorest in SPNs occurring within the first five years after FPN.[4] Risk factors for early SPNs have only been assessed in one previous study which showed radio- and chemotherapy to be associated with SPNs.[4]

Cancer predisposition syndromes (CPSs) are genetic conditions associated with an increased risk of development of neoplasms.[13-15] More than 40 different CPSs have been associated with a wide range of neoplasms during childhood and adolescence.[16] So far, there are no studies that assessed CPSs as possible risk factor for early SPNs on a population-based level. Our study aimed to describe the incidence of SPNs in Swiss childhood cancer patients under 21 years of age, identify CPSs associated with early SPNs, and assess potential risk factors for SPNs with a special focus on CPSs.

## 2. Patients and methods

### 2.1. Study design, study population and inclusion criteria

We performed a cohort study based on the Swiss Childhood Cancer Registry (SCCR). The SCCR collects data on all children and adolescents diagnosed with a neoplasm including leukemias, lymphomas, central nervous system tumors, and malignant solid tumors under the age of 21 years since 1976 in Switzerland.[17] For this analysis, we restricted the diagnosis period to 1986-2015 which has a high completeness of registration.[18] We included all neoplasms fulfilling criteria for the 12 main groups of the International Classification of Childhood Cancer, third edition (ICCC-3),[18] and excluded patients with non-melanoma skin cancer due to incomplete reporting. The Ethics Committee of the Canton of Bern gave its approval to the SCCR (KEK-BE 166/2014).

### 2.2. Demographic, cancer, and treatment characteristics

We extracted demographic, cancer, and treatment characteristics for all childhood cancer patients with an FPN from the SCCR. This included information on sex, age at diagnosis, year of diagnosis, diagnosis classified into 12 main groups of the ICCC-3, [19] relapse, underlying diseases, treatments, and survival. Treatment was categorized into binary variables indicating exposure to chemotherapy, radiotherapy, and hematopoietic stem cell transplantation (HSCT).

### 2.3. Cancer predisposition syndromes

We collected and reviewed medical records from patients for whom underlying diseases were reported to the SCCR database (**Supplementary Text A1**). Yearly follow-up information was continuously reported to the SCCR from hospitals involved in childhood cancer care. We classified underlying diseases as cancer predisposition syndromes (CPSs) if they were reported in the literature to be associated with an increased relative risk of neoplasms compared to the general population.[20]

### 2.4. Identification and ascertainment of second primary neoplasms

We identified SPNs using the SCCR **(Supplementary Text A2)**.[21] We included all SPNs that occurred before age 21 years. We verified all SPNs with the original data source (pathology reports, medical records, cantonal cancer registry and SCCR notification forms, or death certificates) and coded them according to the International Agency for Research on Cancer (IARC) criteria for SPN coding.[22] In brief, we included neoplasms according to the ICCC-3 classification which originated in different tissues or had a different morphology than the FPN and excluded non-melanoma skin cancer, progression, transformation, metastasis, and relapse of FPN.

### 2.5. Statistical analysis

We defined time at risk as the time from date of diagnosis of the FPN until occurrence of the first SPN or (i) last date of hospital follow-up, (ii) last contact with the SCCR (e.g. survivor questionnaire reply or phone calls), or (iii) date of last vital status from cantonal cancer registries, whichever occurred last under 21 years of age. We censored all follow-up information after December 31^st^, 2015, the end of our observation period. To assess mortality, we used data from the SCCR which performs regular updates of survival status in its cohort with the Swiss national death registry.

We stratified observed SPNs by sex, ICCC-3 diagnostic group, age at diagnosis in 5-year groups, and 10-year calendar period of diagnosis. As reference for expected neoplasms, we used incidence rates from the SCCR for ages 0-14 years and from the Swiss Federal Office of Health for the Swiss population for ages 15-20 years[23,24] and multiplied the incidence rate by the observation time for each stratum. We calculated standardized incidence ratios (SIRs) by dividing observed numbers of SPNs in childhood cancer patients by expected numbers in the general population (indirect standardization). We calculated the absolute excess risk (AER) per 100,000 person-years by subtracting the expected number of FPNs from observed number of SPNs, dividing by observation time at risk in person-years, and multiplying by 100,000.

We used competing risk regression models as described by Fine and Gray[25] to estimate hazard ratios (HRs) for pre-defined possible risk factors with death as competing risk to the main outcome. For missing treatment information, we performed multiple imputation by chained equations (MICE) (**Supplementary Text A3**). We then performed univariable competing risk regressions for having an SPN with the following potential risk factors: sex, age at FPN diagnosis, year of FPN diagnosis, diagnostic group of FPN (according to ICCC-3), relapse of FPN, underlying CPSs, and treatment characteristics (chemotherapy, radiotherapy, HSCT as binary variables). We performed a multivariable competing risk regression including sex, and all characteristics associated in the univariable models with a two-sided p-value of ≤0.1. As a sensitivity analysis, we also performed all regression analyses using complete cases only. We plotted the cumulative incidence of SPNs over time since FPN diagnosis with death as competing risk and stratified patients by presence of cancer predisposition syndromes. We used STATA software for statistical analysis (Version 15.1, Stata Corporation, Austin, TX).

## 3. Results

### 3.1. Cohort characteristics

We included 8,074 patients in our study (**Supplementary Figure A4**). Cumulative follow-up time was 47,464 person-years and median attained age at last follow-up was 18.8 years (interquartile range [IQR] 13.1-20.9). Among included patients, 92 patients were diagnosed with one SPN and two patients with two SPNs during follow-up. Median age at FPN diagnosis was 6.8 years (IQR 2.6-14.2) for patients with an SPN and 10.7 years (IQR 4.0-16.5) for those without (**Table 1**). SPNs occurred mainly after leukemias (n=29, 31%), CNS tumors (n=16, 17%), and lymphomas (n=13, 14%), which represent also the most common FPNs in this age range. A higher proportion of patients who later developed SPNs had undergone treatment with chemotherapy, radiotherapy, and/or HSCT for their FPN compared to those who did not develop an SPN. Relapse after FPN was seen in 23% (n=22) in those with and 18% (n=1,451) in those without SPNs. Overall survival up to 21 years of age was lower in patients with SPNs (64%) than in those without (79%).

**Table 1:**
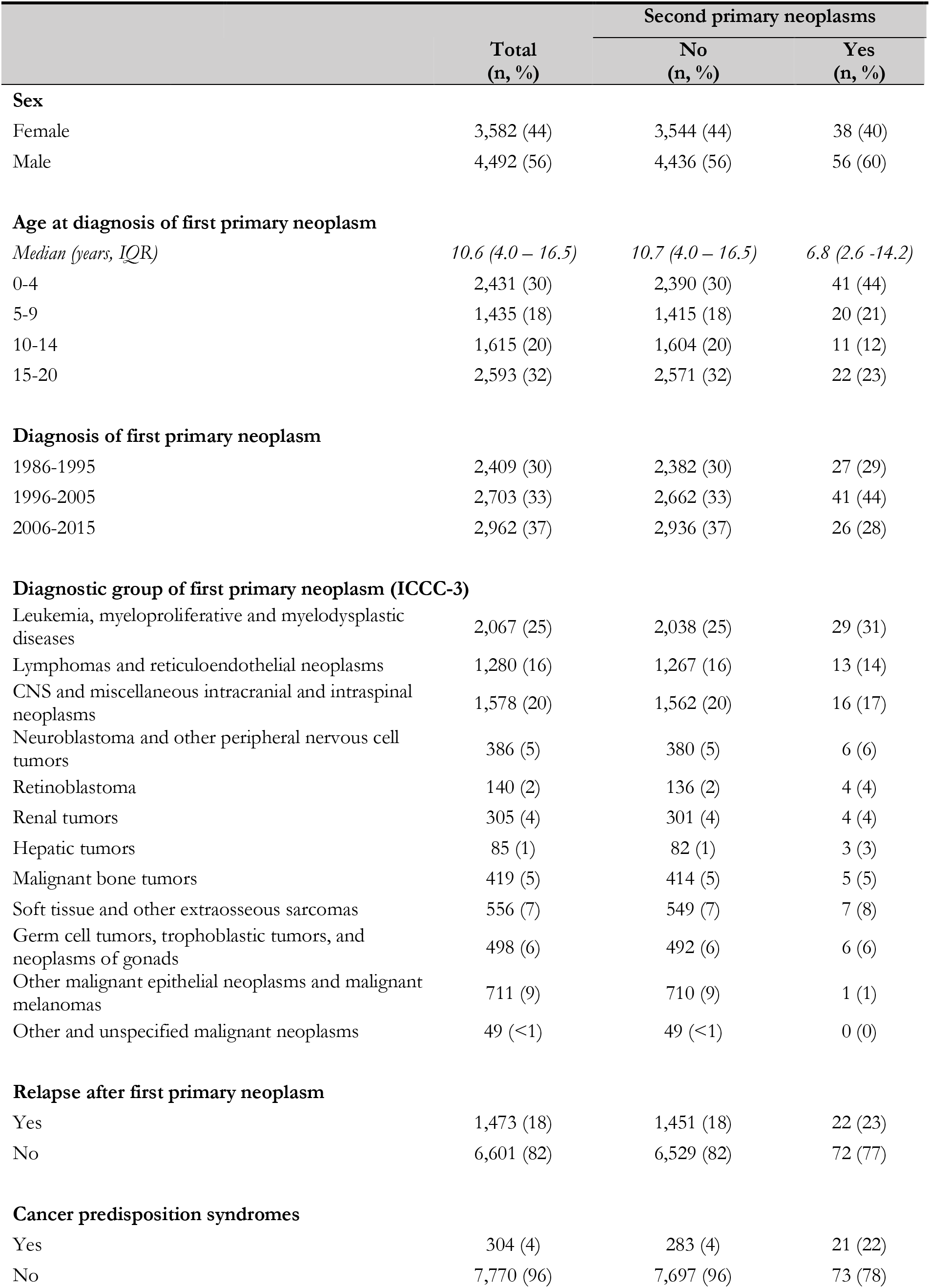
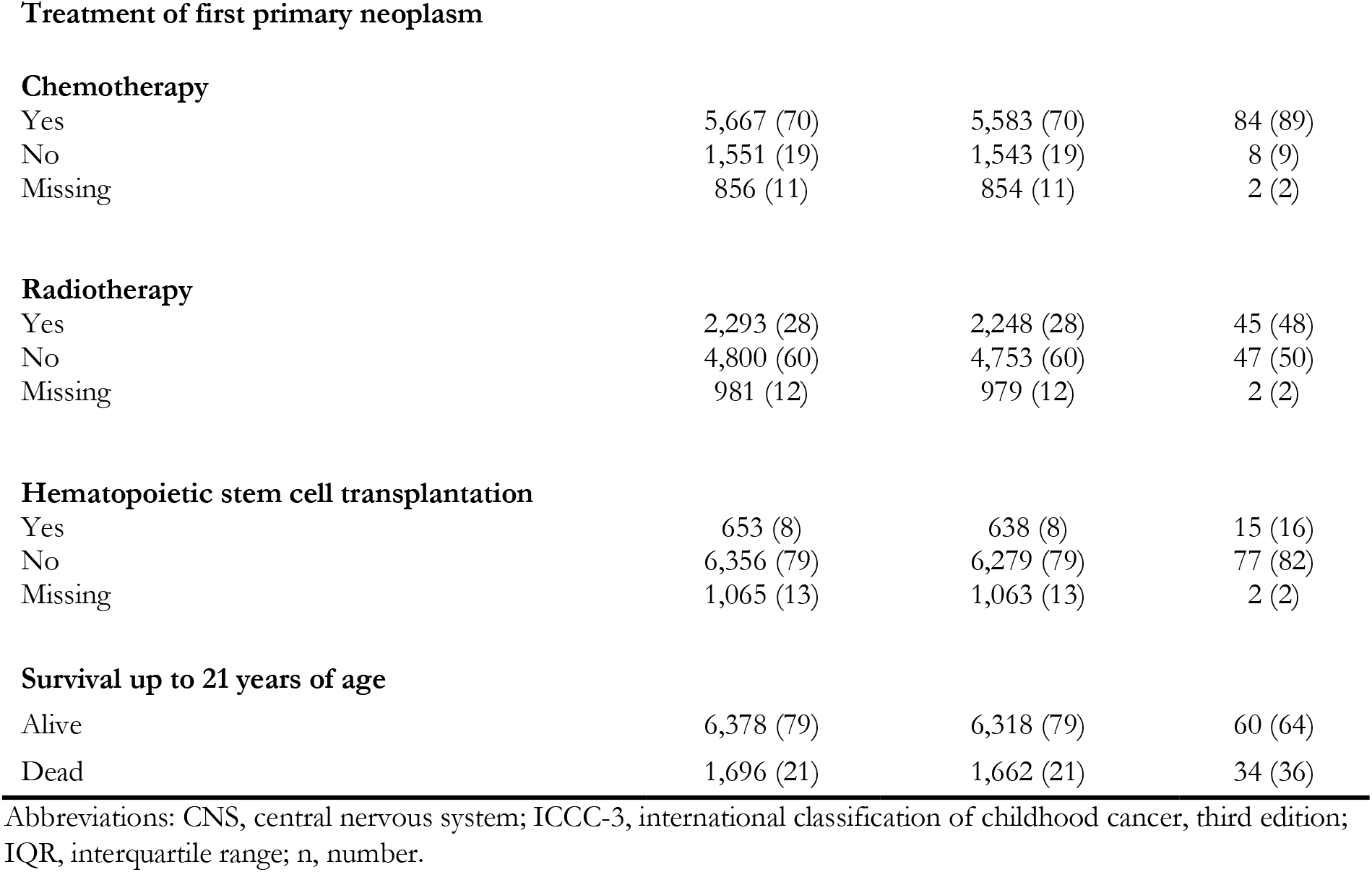
Demographic, cancer, and treatment characteristics of 8,074 Swiss childhood cancer patients diagnosed with a first primary neoplasm between 1986-2015, overall and stratified by diagnosis of second primary neoplasm.

### 3.2. Cancer predisposition syndromes

We identified 304 (4%) childhood cancer patients with a cancer predisposition syndrome (**Supplementary Table A5**). The most common CPSs were neurofibromatosis types 1 and 2 (n=89, 29% of patients with CPSs), trisomy 21 and mosaic trisomies (n=73, 24%), and familial/ bilateral retinoblastoma (n=45, 15%). In total, 27 different CPSs were identified, of which the majority (n=20, 74%) was seen in five or less patients each. Twenty-one of 304 patients with CPSs (7%) developed an early SPN under age 21.

### 3.3. Second primary neoplasms

SPNs were seen within the first month up to 16.8 years after FPN diagnosis (**Table 2)**. Median time from diagnosis of FPN to SPN was 4.0 and 4.9 years and median age at SPN was 13.6 and 15 years in those with and without CPSs, respectively. The most common SPNs were CNS tumors (n=20), leukemias and myelodysplastic syndromes (MDS; n=18), malignant epithelial neoplasms and malignant melanomas (n=18). We cross-tabulated FPNs with SPNs and found that first primary leukemias and MDS were commonly associated with second primary malignant epithelial neoplasms (n=8) and with lymphomas (n=7), lymphomas with malignant epithelial neoplasms (n=7), and CNS tumors with subsequent CNS tumors (n=6) (**Supplementary Table A6**).

**Table 2:** Clinical characteristics of 96 second primary neoplasms in 94 patients, overall and stratified by diagnosis of cancer predisposition syndrome.

| Characteristics | Total<br>(n, %) | Cancer predisposition syndrome |  |
| --- | --- | --- | --- |
|  |  | No<br>(n, %) | Yes<br>(n, %) |
| Diagnostic group of second malignant neoplasm (ICCC-3) † |  |  |  |
| Leukemias, myeloproliferative and myelodysplastic diseases | 18 (19) | 16 (22) | 2 (9) |
| Lymphomas and reticuloendothelial neoplasms | 10 (10) | 9 (12) | 1 (5) |
| CNS and miscellaneous intracranial and intraspinal neoplasms | 20 (21) | 12 (16) | 8 (36) |
| Neuroblastoma and other peripheral nervous cell tumors | 3 (3) | 1 (1) | 2 (9) |
| Retinoblastoma | 0 (0) | 0 (0) | 0 (0) |
| Renal tumors | 5 (5) | 2 (3) | 3 (14) |
| Hepatic tumors | 0 (0) | 0 (0) | 0 (0) |
| Malignant bone tumors | 8 (8) | 5 (7) | 3 (14) |
| Soft tissue and other extraosseous sarcomas | 9 (9) | 7 (9) | 2 (9) |
| Germ cell tumors, trophoblastic tumors, and neoplasms of gonads | 5 (5) | 4 (5) | 1 (5) |
| Other malignant epithelial neoplasms and malignant melanomas* | 18 (19) | 18 (24) | 0 (0) |
| Other and unspecified malignant neoplasms | 0 (0) | 0 (0) | 0 (0) |
| Time from first primary to second primary neoplasm |  |  |  |
| Median (years, IQR) | 4.6 (2.7 – 8.4) | 4.9 (2.7 – 8.4) | 4.0 (2.7 – 8.3) |
| Age at diagnosis of second primary neoplasm † |  |  |  |
| Median (years, IQR) | 14.8 (9.8 - 17.7) | 15.0 (9.9 – 17.7) | 13.6 (7.5 – 17.3) |
| 0-4 years | 6 (6) | 3 (4) | 3 (14) |
| 5-9 years | 20 (21) | 17 (23) | 3 (14) |
| 10-14 years | 24 (25) | 17 (23) | 7 (32) |
| 15-20 years | 46 (48) | 37 (50) | 9 (41) |
| Year of diagnosis of second primary neoplasm † |  |  |  |
| 1986-1995 | 8 (8) | 8 (11) | 0 (0) |
| 1996-2005 | 32 (33) | 29 (39) | 3 (14) |
| 2006-2015 | 56 (58) | 37 (50) | 19 (86) |

Abbreviations: CNS, central nervous system; ICC3, international classification of childhood cancer, third edition; n, number.
\* Other epithelial neoplasms that were identified in this group: thyroid carcinoma, n=10; malignant melanoma, n=4; mucoepidermoid carcinoma of the parotid gland, n=3; adenocarcinoma of the bile ducts, n=1.
† Including multiple SPNs occurring in two patients.

### 3.4. Standardized incidence ratios and absolute excess risks

The risk of having a SPN in childhood cancer patients was more than 10-fold compared to the risk of having a neoplasm in the general population (SIR 10.6; 95%-CI 8.7-13.1), (**Table 3**). We found the highest risk ratios for renal tumors (SIR 24.4, 95%-CI 10.2-58.6), soft tissue sarcomas (SIR 18.5, 95%-CI 9.6-35.6), neuroblastoma (SIR 16.8, 95%-CI 5.2-52.1), and other malignant epithelial neoplasms (SIR 15.4, 95%-CI 9.7-24.5). The AER for any SPN was 179 per 100,000 person-years (py), with the highest AERs in CNS tumors (39/100,000), epithelial neoplasms (36/100,000), and leukemias (30/100,000).

**Table 3:** Standardized incidence ratio and absolute excess risk of second primary neoplasms in childhood cancer patients in relation to the general population stratified by ICCC-3 classification.

| Diagnostic group of second primary neoplasm | Observed*<br>(n) | Expected<br>(n) | SIR (CI) | AER (CI; per 100,000 person-years) |
| --- | --- | --- | --- | --- |
| Leukemia | 16 | 1.8 | 9.1 (5.6 - 14.9) | 30.0 (13.5 - 46.5) |
| Lymphoma | 10 | 1.9 | 5.3 (2.9 - 9.9) | 17.1 (4.1 - 30.2) |
| CNS neoplasms | 20 | 1.4 | 14.8 (9.6 - 22.9) | 39.0 (20.8 - 57.8) |
| Neuroblastoma | 3 | 0.2 | 16.8 (5.2 - 52.1) | 5.9 (-1.2 - 13.1) |
| Retinoblastoma | 0 | 0.1 | 0 | -0.2 (-) |
| Renal tumors | 5 | 0.2 | 24.4 (10.2 - 58.6) | 10.1 (0.9 - 19.4) |
| Hepatic tumors | 0 | 0.1 | 0 | -0.2 (-) |
| Malignant bone tumors | 8 | 0.6 | 13.4 (6.7 - 26.9) | 15.6 (3.9 - 27.3) |
| Soft tissue sarcomas | 9 | 0.5 | 18.5 (9.6 - 35.6) | 17.9 (5.5 - 30.3) |
| Germ cell tumors & neoplasms of gonads | 5 | 0.9 | 5.6 (2.3 - 13.4) | 8.6 (-0.6 - 17.9) |
| Other malignant epithelial neoplasms | 18 | 1.2 | 15.4 (9.7 - 24.5) | 36 (17.9 - 53.0) |
| Other and unspecified malignant neoplasms | 0 | 0.2 | 0 | -0.4 (-) |
| **All categories** | **94** | **8.9** | **10.6 (8.7 - 13.1)** | **179.4 (139.3 - 219.4)** |

Abbreviations: AER, absolute excess risk; CI, 95% confidence interval; CNS, central nervous system; n, number; SIR, standardized incidence ratio.

### 3.5. Predictors for second primary neoplasms

We found that CPSs, chemotherapy, radiotherapy, HSCT, age at FPN diagnosis, and year of FPN diagnosis were associated with SPNs in the unadjusted competing risk analysis (**Table 4**). There was no evidence of association of sex, diagnostic group, and relapse with the development of SPNs. In the multivariable analysis, diagnosis of CPSs (HR 7.8, 95%-CI 4.8-12.7), age at FPN diagnosis 15-20 years (HR 1.9; 95%-CI 1.1-3.2, chemotherapy (HR 2.2; 95%-CI 1.1-4.6), radiotherapy (HR 1.9; 95%-CI 1.2-2.9), and HSCT (HR 1.8, CI 1.0-3.3) remained independent predictors of SPN diagnosis. The hazard ratios were similar in the complete case analysis (**Supplementary Table A7**). Cumulative incidence of SPNs in participants with CPSs was 8% at 10 years, and 23% at 20 years after FPN diagnosis, compared to 1.7% at 10 years and 3% at 20 years in those without CPSs (**Figure 1**).

**Table 4:**
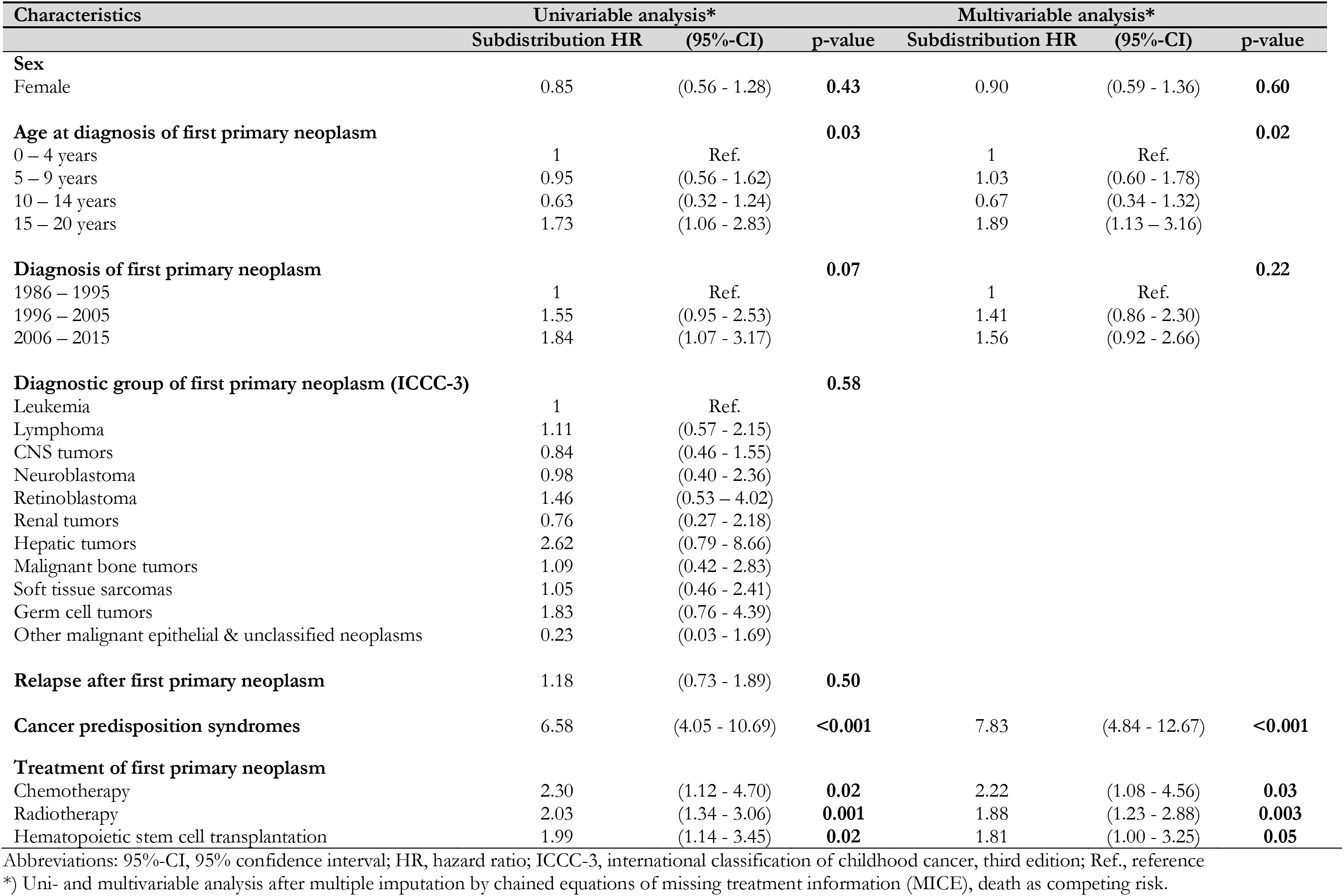
Competing risk regression analysis of demographic, cancer, and treatment characteristics and their association with second neoplasm incidence.

| Characteristics | Univariable analysis* |  |  | Multivariable analysis* |  |  |
| --- | --- | --- | --- | --- | --- | --- |
|  | Subdistribution HR | (95%-CI) | p-value | Subdistribution HR | (95%-CI) | p-value |
| **Sex** |  |  |  |  |  |  |
| Female | 0.85 | (0.56 - 1.28) | **0.43** | 0.90 | (0.59 - 1.36) | **0.60** |
| **Age at diagnosis of first primary neoplasm** |  |  | **0.03** |  |  | **0.02** |
| 0 – 4 years | 1 | Ref. |  | 1 | Ref. |  |
| 5 – 9 years | 0.95 | (0.56 - 1.62) |  | 1.03 | (0.60 - 1.78) |  |
| 10 – 14 years | 0.63 | (0.32 - 1.24) |  | 0.67 | (0.34 - 1.32) |  |
| 15 – 20 years | 1.73 | (1.06 - 2.83) |  | 1.89 | (1.13 - 3.16) |  |
| **Diagnosis of first primary neoplasm** |  |  | **0.07** |  |  | **0.22** |
| 1986 – 1995 | 1 | Ref. |  | 1 | Ref. |  |
| 1996 – 2005 | 1.55 | (0.95 - 2.53) |  | 1.41 | (0.86 - 2.30) |  |
| 2006 – 2015 | 1.84 | (1.07 - 3.17) |  | 1.56 | (0.92 - 2.66) |  |
| **Diagnostic group of first primary neoplasm (ICCC-3)** |  |  | **0.58** |  |  |  |
| Leukemia | 1 | Ref. |  |  |  |  |
| Lymphoma | 1.11 | (0.57 - 2.15) |  |  |  |  |
| CNS tumors | 0.84 | (0.46 - 1.55) |  |  |  |  |
| Neuroblastoma | 0.98 | (0.40 - 2.36) |  |  |  |  |
| Retinoblastoma | 1.46 | (0.53 - 4.02) |  |  |  |  |
| Renal tumors | 0.76 | (0.27 - 2.18) |  |  |  |  |
| Hepatic tumors | 2.62 | (0.79 - 8.66) |  |  |  |  |
| Malignant bone tumors | 1.09 | (0.42 - 2.83) |  |  |  |  |
| Soft tissue sarcomas | 1.05 | (0.46 - 2.41) |  |  |  |  |
| Germ cell tumors | 1.83 | (0.76 - 4.39) |  |  |  |  |
| Other malignant epithelial & unclassified neoplasms | 0.23 | (0.03 - 1.69) |  |  |  |  |
| **Relapse after first primary neoplasm** | 1.18 | (0.73 - 1.89) | **0.50** |  |  |  |
| **Cancer predisposition syndromes** | 6.58 | (4.05 - 10.69) | **<0.001** | 7.83 | (4.84 - 12.67) | **<0.001** |
| **Treatment of first primary neoplasm** |  |  |  |  |  |  |
| Chemotherapy | 2.30 | (1.12 - 4.70) | **0.02** | 2.22 | (1.08 - 4.56) | **0.03** |
| Radiotherapy | 2.03 | (1.34 - 3.06) | **0.001** | 1.88 | (1.23 - 2.88) | **0.003** |
| Hematopoietic stem cell transplantation | 1.99 | (1.14 - 3.45) | **0.02** | 1.81 | (1.00 - 3.25) | **0.05** |

Abbreviations: 95%-CI, 95% confidence interval; HR, hazard ratio; ICC-3, international classification of childhood cancer, third edition; Ref., reference
\*) Uni- and multivariable analysis after multiple imputation by chained equations of missing treatment information (MICE), death as competing risk.

**Figure 1.**
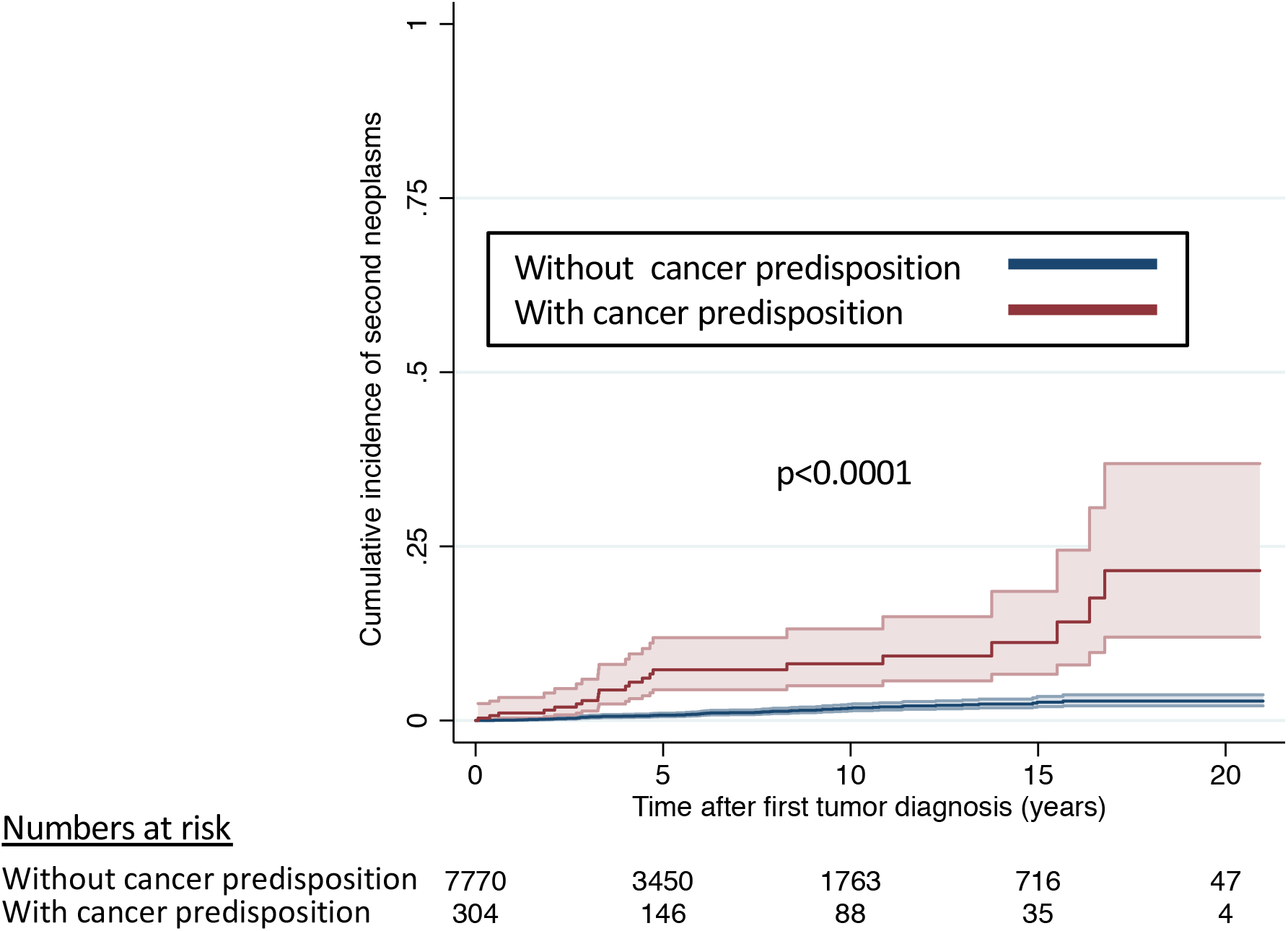
Cumulative incidence of second primary neoplasms with death as competing risk stratified by diagnosis of a cancer predisposition syndrome.

## 4. Discussion

This is the first population-based study to investigate cancer predisposition syndromes with demographic, cancer, and treatment characteristics as risk factors for second primary neoplasms. This study identified CPSs as the most important risk factor for SPNs in pediatric and adolescent patients. The cumulative incidence of SPNs after 20 years was 23% in participants with CPSs and 3% without. Chemotherapy, radiotherapy, HSCT, and older age at FPN diagnosis were also associated with SPNs.

The prevalence of CPSs in our cohort (4%) was lower than the reported prevalence of CPSs in other studies where comprehensive genetic gene panel testing had been performed on all participants.[13,14] A study from St.Jude Children’s Research Hospital assessed 1,120 pediatric cancer patients and found 8.5% affected with CPSs[13] and an international collaborative study in 914 patients identified 7.6% with CPSs.[14] This suggests that many CPSs were not recognized in routine clinical practice in Switzerland.

Our results on the standardized incidence rates and absolute excess risks of second primary neoplasms in childhood cancer patients are comparable with the literature. Previous studies have mostly focused on 3- to 5-year survivors and found SIRs between 5.9 and 11.2.[2,7,9,9,10,26,27] Differences between studies are partially due to sampling variability and differences of follow-up information, but might also be explained by different calendar periods of FPN diagnosis (with diagnoses in the 1990s reporting a lower cumulative incidence than in the 1970s),[7] and different coding of SPNs.[28] Studies from the U.S.,[29] the Nordic countries,[11] Ontario (Canada),[4] and Germany[30] have included SPNs within the first years after diagnosis and found SIRs of 6.5-9.9. Similar to our study, high SIRs for CNS tumors, bone and soft tissue sarcomas were also reported by studies from the U.K. and the Netherlands.[2,27] The absolute excess risk of SPNs in our analysis (AER of 179/100,000py) was comparable with previous reports with AERs of about 110-200/100,000 py.[11,27,29] AERs in the different SPN groups were similar.[4]

We found that cancer predisposition syndromes were the strongest risk factor for early second primary neoplasms. The association of CPSs with SPNs has previously been shown but in a selected cohort of 3,006 5-year survivors from St.Jude Children’s Research Hospital excluding early SMNs. The authors reported an association of CPSs with any SPN in non-irradiated survivors and with selected SPNs (breast cancer and sarcomas) in irradiated survivors.[15]. Similarly to our study, radio- and chemotherapy have been associated repeatedly with SPNs.[2,6,7,27] Older age at FPN diagnosis was previously associated with SPNs in a cohort-based study from Israel on 6,637 5-year survivors[8] and might reflect particular sensitivity of patients to treatment in this age group or variations in neoplasm subtypes and treatments. We found no evidence of an association of relapse of the FPN with occurrence of SPNs. This was in contrast to a previous case-cohort study from the Nordic countries including 196 SPNs which adjusted for radio- and chemotherapy and found particularly second breast cancers associated with relapse of the FPN.[31] The increased toxicity with FPN relapse treatment might only become relevant after longer follow-up. Similarly, female sex was not associated with SPNs in our analysis, which is likely explained by the lack of breast cancers in our young cohort, which was the main SPN in other studies but occurred usually after 21 years of age.[32]

A limitation of our study is, that we were not able to assess which patients had undergone genetic counselling and whether CPSs had been diagnosed by genetic testing. Therefore, it is possible that patients with SPNs had been investigated more often for the presence of CPSs than those without. Still, 22% of patients with early SPNs also being diagnosed with CPSs exceeds what we would expect for the childhood cancer population of about 8%.[13,14] Another limitation is the crude information on treatment exposures which did not allow to relate specific treatment agents to SPNs. Finally, due to the limited number of events in our population, confidence intervals and estimation of incidence ratios and excess risks are wide. A strength of our study is the inclusion of SPNs since diagnosis of FPN which provides a complete assessment of SPNs in the pediatric and adolescent age range. These patients were assessed for SPNs in only few previous studies. Second, due to the continuous collection of information into the SCCR with yearly follow-up information and medical reports, we were able to assemble detailed clinical data from the initial cancer treatment period and from follow-up appointments. Third, our study is based on a nationwide, population-based cancer registry, so that our results are representative for all Switzerland and childhood cancer in countries with access to modern workup and treatment modalities.

Identification of patients at increased risk of second primary neoplasms is imperative to tailor cancer surveillance. The strong association of CPSs with SPNs in our study and occurrence of SPNs within the first years after FPNs in some patients highlight the importance to identify CPSs in childhood cancer patients. Adapted cancer surveillance protocols for specific CPSs include multimodal screening methods[33-35] and yielded survival advantages.[36,37] This is relevant for newly diagnosed patients but also those who have been cured of their primary disease, as the identification of CPSs may have been missed during initial workup and treatment. Appropriate surveillance of patients with CPSs is important and might improve early detection of SPNs and ultimately long-term survival.

### Author contribution statement

Nicolas Waespe: Conceptualization, Methodology, Data preparation and curation, Statistical analysis, Writing - all stages, Visualization; Fabien Belle: Writing - Reviewing and Editing; Shelagh Redmond: Data collection, preparation and curation, Writing - Reviewing and editing; Christina Schindera: Writing - Reviewing and editing; Ben Spycher: Statistical analysis, Writing - Reviewing and Editing; Claudia E. Kuehni: Supervision, Conceptualization, Methodology, Statistical analysis, Writing - Reviewing and Editing. Marc Ansari & Jochen Roessler: Writing - Reviewing and Editing.

## Data Availability

Childhood cancer is a rare disease and to avoid identification of individual patients, data is not available online. The data that support the findings of this study are available upon reasonable request. All data requests should be communicated to the corresponding author at the Institute of Social and Preventive Medicine, University of Bern, Switzerland.

## Acknowledgements

We thank all childhood cancer patients and families for participating in this study. We thank the study team at the Institute of Social and Preventive Medicine, University of Bern (Luzius Mader, Maria Otth, Sven Strebel, Jana Remlinger, Grit Sommer), the data managers of the Swiss Pediatric Oncology Group (Claudia Althaus, Nadine Assbichler, Pamela Balestra, Heike Baumeler, Nadine Beusch, Sarah Blanc, Pierluigi Brazzola, Susann Drerup, Janine Garibay, Franziska Hochreutener, Monika Imbach, Friedgard Julmy, Eléna Lemmel, Rodolfo Lo Piccolo, Heike Markiewicz, Veneranda Mattiello, Annette Reinberg, Renate Siegenthaler, Astrid Schiltknecht, Beate Schwenke, and Verena Stahel) and the data managers and administrative staff at the Institute of Social and Preventive Medicine, University of Bern (Meltem Altun, Erika Brantschen, Katharina Flandera, Elisabeth Kiraly, Verena Pfeiffer, Julia Ruppel, Ursina Roder, and Nadine Lötscher). We thank the Federal Office of Public Health (FOPH) and the National Institute of Cancer Epidemiology and Registration (www.nicer.org) for providing information on cancer incidence rates in Switzerland.This study was supported by the CANSEARCH Foundation, the Swiss National Science Foundation (31BL30_185396), Swiss Cancer Research and the Swiss Cancer League (KLS-3886-02-2016 and KLS/KFS-4825-01-2019, KFS-4722-02-2019), “Stiftung für krebskranke Kinder Regio Basiliensis.” The work of the Swiss Childhood Cancer Registry is supported by the Swiss Pediatric Oncology Group (www.spog.ch), Schweizerische Konferenz der kantonalen Gesundheitsdirektorinnen und -direktoren (www.gdk-cds.ch), Swiss Cancer Research (www.krebsforschung.ch), and Kinderkrebshilfe Schweiz (www.kinderkrebshilfe.ch).

**Abbreviation table**
AER: Absolute excess risk
CI: Confidence Interval
CNS: Central nervous system
CPS: Cancer predisposition syndrome
FPN: First primary neoplasm
HR: Hazard ratio
HSCT: Hematopoietic stem cell transplantation
IARC: International Agency for Research on Cancer
ICCC-3: International Classification of Childhood Cancer, third edition
IQR: Interquartile range
PY: Person-years
SIR: Standardized incidence ratio
SPN: Second primary neoplasm
SCCR: Swiss Childhood Cancer Registry
SPOG: Swiss Pediatric Oncology Group

---

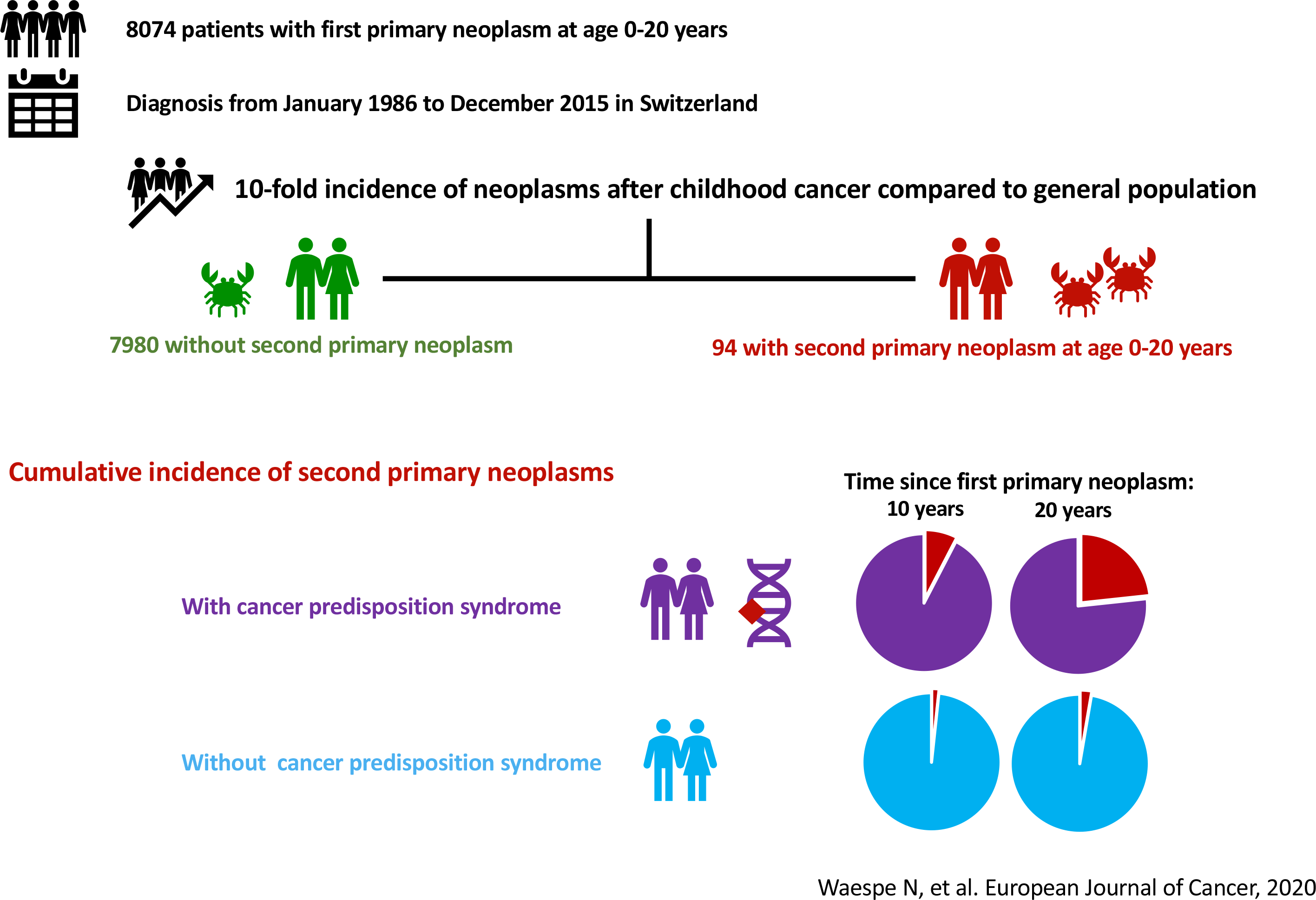

